# Traditional Healing Practices as a Complement or Barrier to Modern Orthopedic Care in White Nile State, Sudan 2024: A cross sectional study

**DOI:** 10.64898/2026.02.10.26346052

**Authors:** Ahmed Mohamed Ahmed Ali, Israa Ismael Zakarya Ismael, Ahmed Elfatih Hasan Hamad, Aisha Ibrahim Ahmd Omer

## Abstract

**Introduction:** Traditional bone-setting remains a culturally significant healthcare practice in low- and middle-income countries, particularly in regions like Sudan where modern orthopedic services are often inaccessible or unaffordable .This study examines the role of traditional healing practices in orthopedic care in White Nile State, Sudan, assessing patient perceptions, treatment effectiveness, and sociocultural factors influencing healthcare choices.

**Methods:** A cross-sectional analytical study was conducted among 147 patients, 7 traditional healers, and 4 orthopedic practitioners in urban and rural areas of White Nile State. Data were collected using structured questionnaires and interviews, focusing on treatment preferences, perceived effectiveness, and barriers to integration. Descriptive and inferential statistics were used to analyze quantitative data, while thematic analysis was applied to qualitative responses from healers and practitioners.

**Results:** Fractures (45.6%) and arthritis (23.1%) were the most common orthopedic conditions. 30.6% of patients initially sought traditional treatment, all eventually utilized modern care (medication 71.4%, surgery 42.9%). Traditional healing was perceived as somewhat effective by 40% of users, whereas 59.9% rated modern care as very effective. Key factors influencing treatment choices included cultural beliefs (29.9%), accessibility (18.4%), and cost (16.3%). No significant demographic associations were found with treatment preference or effectiveness (p > 0.05). Traditional healers predominantly treated dislocations (100%) and fractures (71.4%) using manual techniques, with 57.1% referring complex cases to modern practitioners. Barriers to collaboration included lack of communication (85.7% of healers) and differing treatment philosophies (50% of practitioners).

**Conclusion:** This study highlights the persistent dual reliance on traditional and modern orthopedic care in Sudan, with modern treatments perceived as more effective yet traditional methods remaining culturally entrenched especially in rural areas. The path forward requires bridging these systems through mutual respect, shared protocols, and community engagement to ensure safe, equitable, and effective musculoskeletal care for all Sudanese patients.

## INTRODUCTION

Traditional bone setting (TBS) is a branch of traditional medicine which has deep roots in many countries across the world, although the art, practice and name may differ from region to region. In some regions of the world, TBS has and continuous to be the mainstay or alternative health care option for the population(1). It is a specialized type of traditional medicine that deals with joint manipulation and musculoskeletal injury management .In Sudan, traditional healing practices hold particular cultural significance, with bone-setters continuing to serve as primary care providers for musculoskeletal injuries, especially in rural areas (2). The persistence of this practice reflects complex socio-cultural dynamics, where traditional healers remain more accessible, affordable, and culturally congruent than modern medical services. Some progressive bone-setters have begun incorporating modern elements like X-rays and electrotherapy machines into their practice, yet their fundamental techniques often lack scientific validation. The consequences are frequently severe, with complication rates reaching 20.6% in some studies, creating a significant public health challenge (3).

There might be stories of patients treated successfully by TBSs, but those who seek orthopedic treatment after consulting TBSs often do so due to complications resulting from TBS mismanagement. These complications range from minor limb discrepancies (caused by malunion of fractures) with minimal effect on function to major complications like limb gangrene and death (4). The practice is usually preserved as a family tradition, often passed from father to son or learned through apprenticeship based on experience and spiritual intuition. This method of knowledge transfer promotes the inheritance of improper knowledge of disease prevention and control measures. Unfortunately, studies and research regarding these problems are scarce. A traditional bonesetter is a lay practitioner of bone manipulation who, according to the views of patrons and communities, is skilled in restoring broken bones to full functionality. However, Agrawal defines the modern-day traditional bonesetter as “the unqualified practitioner who takes up the practice of healing without having had any formal training in accepted medical procedures” (5).

Traditional healing practices remain prevalent in Sudan, where modern healthcare services, especially specialized orthopedic care, may not be readily accessible or affordable to many. Delayed or inappropriate orthopedic interventions due to reliance on traditional healing methods may lead to adverse clinical outcomes, such as malunion of fractures or untreated infections. In White Nile State where traditional practices are deeply rooted, it is crucial for modern healthcare systems to be culturally sensitive. Our study in White Nile State investigates the prevalence of traditional healing for orthopedic conditions, compares patient perceptions of traditional versus modern treatments, and analyzes the socio-cultural factors influencing healthcare choices. The findings aim to inform policies that might integrate the strengths of traditional medicine with evidence-based orthopedic care, potentially reducing complications while respecting cultural traditions. As younger generations show declining interest in traditional healing (6), and with persistent gaps in standardization and knowledge transmission, this research addresses urgent questions about the future role of traditional bone-setting in contemporary healthcare systems.

## METHODS

### Study Design and Setting

This study employed a cross sectional analytical design to investigate the role of traditional healing practices as either a complement to or a barrier to modern orthopedic care in White Nile State, Sudan. The state consists of eight localities, and participants in this study—including patients and orthopedic specialists— were selected from the four largest hospitals located in the major localities: Rabak, Kosti, Aljabaleen, and Aldweem. Traditional healers who took part in the study were also from rural areas within these same localities. The research was conducted in both urban and rural areas to ensure representation of different healthcare seeking behaviors.

### Study Population and Sampling

The study included three key participant groups: patients diagnosed with orthopedic conditions, traditional healers providing orthopedic-related treatments, and modern orthopedic practitioners working in hospitals and clinics within White Nile State. Systematic random sampling was used to select the samples while sample size was calculated using the sample size of unknown population (Cochran’s formula).

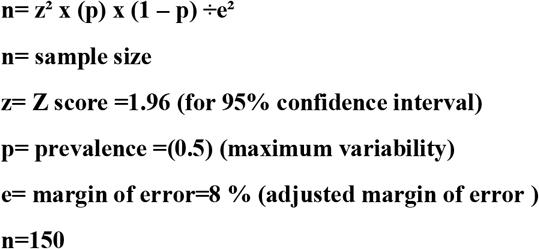

A total of 147 patients were recruited using a convenience sampling approach from healthcare facilities. This ensured that participants represented diverse experiences with both modern and traditional treatment modalities. In addition to patient participants, the study totally covered traditional healers and orthopedic practitioners despite the small sample size. Seven traditional healers, all actively practicing in White Nile State, were selected to provide insights into their methods, treatment philosophies, and interactions with modern medical systems. Likewise, four orthopedic practitioners were included, representing different levels of experience in treating orthopedic conditions and managing patients who had previously sought traditional healing. While the number of traditional healers and orthopedic practitioners was limited, their responses provided a thorough understanding of the intersection between modern and traditional care, ensuring a well-rounded perspective on the research topic.

### Data Collection and Instrumentation

Data were collected by the authors and well-trained medical practitioner using a scientifically validated, self-structured questionnaire developed through an extensive review of existing literature on traditional healing and modern orthopedic care. The questionnaire was developed by the authors specifically for this study. The questionnaire was developed by the authors specifically for this study, and Its quality was assured by a panel of experts in orthopedic medicine and public health through the application of high standards and strategies.

The questionnaire for patients was divided into several sections. The first section collected demographic data, including age, gender, education level, residency, and occupation. The second section focused on orthopedic conditions, asking participants about their diagnoses, duration of illness, and types of treatments sought. The third section explored traditional healing practices, including specific methods used (such as herbal remedies, massage, or spiritual healing) and patient perceptions of effectiveness. The fourth section covered modern orthopedic treatments, detailing the types of care received (such as medication, surgery, or physical therapy) and perceived effectiveness. Additional sections examined the challenges and benefits of using both treatment modalities, factors influencing treatment decisions (cost, accessibility, cultural beliefs, recommendations), and patients’ willingness to integrate traditional healing with modern orthopedic care.

For traditional healers, structured interviews were conducted to gather information on their years of experience, commonly treated orthopedic conditions, treatment methods, and effectiveness assessment criteria. The interviews also explored their referral practices to modern healthcare providers, perceptions of modern orthopedic care, and potential barriers to collaboration.

Similarly, orthopedic practitioners were interviewed using a structured guide designed to capture their awareness of patient use of traditional healing, the frequency of such practices, challenges encountered when managing patients who had previously undergone traditional treatments, and their perspectives on the integration of traditional and modern orthopedic care. The interviews also explored barriers to collaboration, including cultural differences, communication gaps, and differences in treatment philosophies.

### Data Analysis

This study used the Statistical Package for Social Sciences (SPSS26) to analyse the collected data. Descriptive statistics, such as frequencies and percentages, were calculated for all variables in the questionnaire. Chi-squared tests were then employed to explore the relationships between various factors, including treatment preferences, and effectiveness ratings and demographic characteristics. A statistical significance level of p < 0.05 was used to determine the significance of these associations.

## RESULTS

The demographic characteristics of the participants were as follow: The majority of participants were male (60.5%), with (33.3%) aged 6-27 years, 29.3% aged 28-49 years, and 37.4% aged 50 years or older. In terms of education, 29.9% had no formal education, 29.9% had primary education, 17.7% had secondary education, and 22.4% had higher education. A slight majority of participants resided in rural areas (53.1%), and the most common occupation was free worker (32.7%), followed by unemployed (36.1%), student (19.0%), and employee (12.2%).

(Table 1) presents the prevalence of orthopedic conditions and treatment patterns among patients. The most common conditions were fractures (45.6%), arthritis (23.1%), and dislocations (8.2%). The majority of participants (53.7%) had experienced their condition for less than 6 months. While 30.6% sought traditional treatment, all participants (100%) sought modern orthopedic care. Among those who sought traditional treatment, the most common method was massage or manipulation (91.1%), followed by herbal remedies and Hijama (4.4% each). Modern treatments included medication (71.4%), surgery (42.9%), physical therapy (10.9%), and fixation by splint (5.4%).

**Table 1:** Prevalence of Orthopedic Conditions and Patterns of Traditional vs. Modern Treatment among Patients (n=147)

| Variable | N(%) |
| --- | --- |
| **Diagnosed orthopedic condition** |  |
| Arthritis | 34 (23.1%) |
| Bone deformities | 4 (2.7%) |
| Dislocations | 12 (8.2%) |
| Fractures | 67 (45.6%) |
| Skeletal pain | 13 (8.9%) |
| Felon finger | 1 (0.7%) |
| Joint swelling | 2 (1.4%) |
| Muscle tearing | 8 (5.4%) |
| Osteomalacia | 1 (0.7%) |
| Osteomyelitis | 1 (0.7%) |
| Osteoporosis | 1 (0.7%) |
| Sciatica | 1 (0.7%) |
| Tendon rupture | 2 (1.4%) |
| **Duration of the condition** |  |
| Less than 6 months | 79 (53.7%) |
| 6 months to 1 year | 18 (12.2%) |
| More than 1 year | 50 (34.0%) |
| **Sought traditional treatment** |  |
| Yes | 45 (30.6%) |
| No | 102 (69.4%) |
| **Traditional healing methods received** |  |
| Herbal remedies | 2 (4.4%) |
| Massage or manipulation (Traditional splint) | 41 (91.1%) |
| Hijama | 2 (4.4%) |
| **Sought modern orthopedic care** |  |
| Yes | 147 (100.0%) |
| **Modern treatments received (Check all that apply)** |  |
| Medication | 105 (71.4%) |
| Physical therapy | 16 (10.9%) |
| Surgery | 63 (42.9%) |
| Fixation by splint | 8 (5.4%) |

Regarding the utilization of traditional healer treatment. No statistically significant associations were found between age, gender, education level, residency, or occupation and the use of traditional treatment (all p-values > 0.05). Among the 45 participants who sought traditional healing, the majority were male (71.1%). In terms of age distribution, 44.4% were aged 6-27 years, 17.8% were aged 28-49 years, and 37.8% were 50 years or older. Regarding education, 24.4% had no formal education, 33.3% had primary education, 22.2% had secondary education, and 20.0% had higher education. A majority resided in rural areas (60.0%), and the occupational breakdown included employees (11.1%), unemployed individuals (24.4%), students (28.9%), and free workers (35.6%).

The result about the practices of traditional healers indicate that the majority (42.9%) had 5-10 years of experience, and all treated dislocations (100%), while 71.4% treated fractures. The primary method used was massage or manipulation (100%). On the other hand the experience of orthopedic practitioners. Most (75.0%) had less than 5 years of experience, and the most commonly treated conditions were fractures (75.0%), arthritis (50.0%), and dislocations (50.0%).

As in table (5) the results explores collaboration between modern and traditional practices. None of the practitioners had collaborated with traditional healers, and barriers included cultural barriers (25.0%), different treatment philosophies (50.0%), and lack of communication (25.0%). Strategies for integration included developing joint treatment protocols (25.0%) and promoting mutual respect (50.0%). Only 25.0% supported formal integration, suggesting direct contact and communication as steps forward.

## Discussion

Giving the rising number of musculoskeletal injuries in Sudan, and the continued use of traditional bone setters (TBS) even though modern orthopedic care is available, it is important to understand why many patients use both systems. This study helps explain how cultural beliefs and treatment outcomes are connected, and gives insight into why people rely on both traditional and modern healthcare.

This study shows that traditional healing in White Nile State, Sudan, can play both a complement and barrier role in modern orthopedic care. Many patients, especially those in rural areas, prefer traditional methods like massage and splinting because they are cheaper, easier to reach, and match their cultural beliefs (Table 1). These methods are often used as the first treatment for bone and joint problems. While some patients felt they were helpful, many experienced complications like poor healing or delays, which later led them to seek hospital treatment (Table 2). This means traditional healing can support modern care if used early and safely, but it can also be a problem when it causes delays in proper medical treatment.

**Table 2:** Effectiveness and factors influencing both traditional and modern orthopedic care among Patients (n=147)

| Variable | N(%) |
| --- | --- |
| **Effectiveness of traditional healing methods (n=45)** |  |
| Very effective | 4 (8.9%) |
| Somewhat effective | 18 (40.0%) |
| Unsure | 5 (11.1%) |
| Not effective | 18 (40.0%) |
| **Effectiveness of modern orthopedic treatment** |  |
| Very effective | 88 (59.9%) |
| Somewhat effective | 37(25.1%) |
| Unsure | 12 (8.2%) |
| Not effective | 10 (6.8%) |
| **Challenges or benefits from using both traditional and modern treatments (n=45)** |  |
| Both Traditional healing and Modern care improve outcome | 8 (17.7%) |
| Modern care leads to complications, while Traditional healing improves outcome | 1 (2.2%) |
| Traditional healing leads to complications, while modern care improves outcome | 10(22.2%) |
| None | 26(57.8%) |
| **Factors influencing treatment choice (Check all that apply)** |  |
| Cost | 24(16.3%) |
| Accessibility | 27(18.4%) |
| Cultural beliefs | 44(29.9%) |
| Recommendations from family or friends | 26(17.7%) |
| Perceived effectiveness | 36(24.5%) |
| **Interest in integrated approach** |  |
| Yes | 41(27.9%) |
| Unsure | 67(45.6%) |
| No | 39(26.5%) |

In our study, it was obvious that demographic factors like age, gender, education level, residency, and occupation were not strongly linked to whether people used traditional healers. This result is different from some international studies that found higher use of traditional medicine among women or people with lower education ^(7)^. However, it supports other research showing that cultural beliefs and social traditions often have a stronger impact on healthcare choices than income or education level, especially in low- and middle-income countries (LMICs) ^(8)^. The higher number of male (71.1%) and rural (60.0%) participants who used traditional healing in our study (Table 4) matches a common pattern in the region, where these practices remain common regardless of background ^(9)^.

Although we didn’t find a big difference between rural and urban participants, studies from other areas like West Africa show more use of traditional healing in rural communities, likely because modern healthcare is harder to access. In our study, most participants said they turned to traditional healers because of cultural trust and spiritual beliefs (Table 2). This is different from findings in where people often choose traditional bone-setting because they believe it leads to faster recovery ^(10)^.

We also found no strong link between patient backgrounds and the perceived effectiveness of the traditional healers (Table 3). This supports other studies in LMICs, showing that people rely on traditional healing mainly because of belief and trust, not their age, income, or education ^(11)^.

**Table 3.** shows association between demographic characteristics and sought traditional healer treatment (n=147).

| Variable | Traditional treatment |  |  | P value |
| --- | --- | --- | --- | --- |
|  | Yes (N=45) | No (N=102) | Total (N=147) |  |
| **Age group** |  |  |  |  |
| 6-27 years | 20.0 (44.4%) | 29.0 (28.4%) | 49.0 (33.3%) | 0.070 |
| 28-49 years | 8.0 (17.8%) | 35.0 (34.3%) | 43.0 (29.3%) |  |
| more or equal 50years | 17.0 (37.8%) | 38.0 (37.3%) | 55.0 (37.4%) |  |
| **Gender** |  |  |  |  |
| Male | 32.0 (71.1%) | 57.0 (55.9%) | 89.0 (60.5%) | 0.082 |
| Female | 13.0 (28.9%) | 45.0 (44.1%) | 58.0 (39.5%) |  |
| **Education Level:** |  |  |  |  |
| No formal education | 11.0 (24.4%) | 33.0 (32.4%) | 44.0 (29.9%) | 0.608 |
| Primary | 15.0 (33.3%) | 29.0 (28.4%) | 44.0 (29.9%) |  |
| Secondary | 10.0 (22.2%) | 16.0 (15.7%) | 26.0 (17.7%) |  |
| Higher education | 9.0 (20.0%) | 24.0 (23.5%) | 33.0 (22.4%) |  |
| **Residency** |  |  |  |  |
| Rural | 27.0 (60.0%) | 51.0 (50.0%) | 78.0 (53.1%) | 0.263 |
| Urban | 18.0 (40.0%) | 51.0 (50.0%) | 69.0 (46.9%) |  |
| **occupation** |  |  |  |  |
| employee | 5.0 (11.1%) | 13.0 (12.7%) | 18.0 (12.2%) | 0.114 |
| unemployed | 11.0 (24.4%) | 42.0 (41.2%) | 53.0 (36.1%) |  |
| student | 13.0 (28.9%) | 15.0 (14.7%) | 28.0 (19.0%) |  |
| free worker | 16.0 (35.6%) | 32.0 (31.4%) | 48.0 (32.7%) |  |

Modern orthopedic treatment was rated as “very effective” by 59.9% of participants, which is similar to findings in other countries that show success with surgical and evidence-based treatments. However, some patients were not fully satisfied—25.1% said it was “somewhat effective” and 6.8% said it was “not effective” (Table 4). This matches reports from LMICs where long waiting times, high costs, and lack of personal care make people less satisfied. In Sudan as seen in Ghana these challenges often lead patients to try traditional care first, only going to hospitals after problems appear (^12, 13^).

**Table 4:** shows association of demographic characteristics with effectiveness of modern care medicine among patients (n=147)

| Variable | Effectiveness of modern care medicine |  |  | P value |
| --- | --- | --- | --- | --- |
|  | effective (N=125) | Unsure (N=12) | Not effective (N=10) |  |
| **Age** |  |  |  |  |
| Mean (SD) | 41.4 (21.3) | 28.1 (22.4) | 35.3 (18.5) | 0.129 |
| Range | 1.0 - 110.0 | 1.0 - 70.0 | 11.0 - 66.0 |  |
| **Gender** |  |  |  |  |
| Female | 50.0 (40.0%) | 6.0 (50.0%) | 3.0 (30.0%) | 0.659 |
| Male | 75.0 (60.0%) | 6.0 (50.0%) | 7.0 (70.0%) |  |
| **Occupation** |  |  |  |  |
| Employee | 16.0 (12.8%) | 2.0 (16.7%) | 0.0 (0.0%) | 0.327 |
| free worker | 39.0 (31.2%) | 2.0 (16.7%) | 6.0 (60.0%) |  |
| Student | 24.0 (19.2%) | 3.0 (25.0%) | 1.0 (10.0%) |  |
| Unemployed | 46.0 (36.8%) | 5.0 (41.6%) | 3.0 (30.0%) |  |
| **Education Level:** |  |  |  |  |
| Higher education | 30.0 (24.0%) | 2.0 (16.7%) | 0.0 (0.0%) | 0.463 |
| No formal education | 37.0 (29.6%) | 3.0 (25.0%) | 4.0 (40.0%) |  |
| Primary | 36.0 (28.8%) | 3.0 (25.0%) | 5.0 (50.0%) |  |
| Secondary | 22.0 (17.6%) | 4 .0 (33.3%) | 1.0 (10.0%) |  |
| **Residency** |  |  |  |  |
| Rural | 66.0 (52.8%) | 5.0 (41.6%) | 6.0 (60.0%) | 0.652 |
| Urban | 59.0 (47.2%) | 7.0 (58.3%) | 4.0 (40.0%) |  |

Satisfaction with traditional bone setters (TBS) remains high in Sudan, despite that traditional medicine users in our study reporting inefficacy by 40%. Similar studies in Turkey show over 70% satisfaction, often because the care feels more personal, familiar, and spiritually meaningful. In Nigeria, 85% of people with fractures first visit a TBS, leading to many avoidable problems when they finally go to the hospital. This same trend appears in Sudan and shows the need to reduce risks (^14, 15^).

We also found that fractures (45.6%) and arthritis (23.1%) were the most common conditions among participants (Table 2), reflecting a high burden of musculoskeletal diseases. This is similar to patterns across Africa, where older people face more fragility fractures and younger people suffer trauma-related injuries, especially from road traffic accidents ^(16)^. About 30.6% of our participants used traditional healing—mostly massage or splinting (91.1%)—while all participants also used modern orthopedic care (Table 1). This shows a clear dual reliance on both systems, which is common in many African countries ^(17)^. In Sudan, this is made worse by shortages of healthcare workers, unequal access to services, and the high cost or low quality of modern care.

One of the main issues found in this study is the poor connection between traditional healers and orthopedic doctors. None of the four doctors had worked with traditional healers and said that cultural differences (25.0%), different ways of treating patients (50.0%), and poor communication (25.0%) were the main reasons (Table 6). On the other hand, most of the traditional healers (85.7%) were open to working together, through better communication and shared training (Table 5), which is similar to what has been seen in other low- and middle-income countries ^(18)^. Doctors were more cautious because they were concerned about patient safety, lack of clear rules, and the use of methods not based on science ^(19)^. Still, the willingness of traditional healers to cooperate—like in successful programs in Burkina Faso and Mozambique—shows that working together is possible. Many traditional healers said that for good teamwork to happen, there needs to be mutual respect, proper training, and a clear system for referring patients ^(20)^. If these steps are taken, traditional healing could be safely included to help improve orthopedic care in Sudan.

**Table 5:** Barriers and Collaboration Between Traditional and Modern Orthopedic Practitioners(n=7)

| Variable | N(%) |
| --- | --- |
| **Previous collaboration with modern orthopedic practitioners** |  |
| Made splints for hospital patients and worked with a doctor. | 1(14.3%) |
| No | 6 (85.7%) |
| **Challenges with combined treatment** |  |
| Lack of communication | 6 (85.7%) |
| Cultural differences | 1 (14.3%) |
| **Suggestions for better integration** |  |
| Improved communication and collaboration | 6 (85.7%) |
| Joint training sessions | 1 (14.3%) |

**Table 6:** Perception and Outcomes of Collaboration Between Modern Orthopedic Care and Traditional Healing Practices (n=4)

| Variable | N(%) |
| --- | --- |
| **Collaboration with traditional healers** |  |
| No | 4(100.0%) |
| **Barriers to collaboration** |  |
| Cultural barriers | 1 (25.0%) |
| Different treatment philosophies | 2 (50.0%) |
| Lack of communication | 1 (25.0%) |
| **Integration strategies for patient benefit** |  |
| Developing joint treatment protocols | 1 (25.0%) |
| Promoting mutual respect and understanding | 2 (50.0%) |
| No idea | 1 (25.0%) |
| **Support for formal integration** |  |
| Yes | 1 (25.0%) |
| No | 3 (75.0%) |
| **If the answer is yes, what steps should be taken?** |  |
| Direct contact and found channel for communication | 1(100.0%) |

## Limitation

While this study offers important insights into traditional and modern orthopedic care in Sudan, several limitations should be noted. The mixed-methods approach—combining quantitative patient data (n = 147) with qualitative interviews of traditional healers (n = 7) and orthopedic specialists (n = 4)—strengthened depth but limited statistical power for subgroup comparisons. Although the sample included diverse age groups, education levels, and rural/urban residents (53.1% rural), geographic restriction to White Nile State may reduce generalizability to other regions. The cross-sectional design also prevents assessment of long-term treatment outcomes. Additionally, self-reported effectiveness measures could be influenced by recall or social desirability bias, particularly regarding traditional healing practices. These constraints highlight opportunities for future research with larger, multi-regional samples and longitudinal follow-up.

## Conclusion

This study highlights the persistent dual reliance on traditional and modern orthopedic care in Sudan, with modern treatments perceived as more effective yet traditional methods remaining culturally entrenched especially in rural areas. The path forward requires bridging these systems through mutual respect, shared protocols, and community engagement to ensure safe, equitable, and effective musculoskeletal care for all Sudanese patients.

## Recommendations

Establishing regulated training programs for traditional bone-setters should be prioritized, focusing on basic musculoskeletal anatomy and fracture management. Additionally developing formal referral protocols would help bridge the gap between traditional and modern practitioners, particularly for complex cases where delayed presentation leads to worse outcomes . Also community education campaigns are needed to address persistent misconceptions about modern orthopedic care while respecting cultural traditions.

## Data Availability

All data produced in the present study are available upon reasonable request to the authors

## DECLARATIONS

### Ethics approval and consent to participate

According to the Declaration of Helsinki, ethical approval was obtained from the Health Research Ethical Committee of the Ministry of Health, White Nile State. Additional ethical clearance was also granted by the Community Medicine Department at Al-Zaeem Al-Azhary University. Participants were approached in their tents and were required to provide informed consent before taking part in the study.

For participants with no formal education, the study’s purpose and procedures were explained verbally in a language they understood, and verbal consent was obtained in the presence of a witness before written consent was taken either by signature or thumbprint.

For participants under the age of 16, informed consent was obtained from their parents or legal guardians.

### Consent for publication

Not applicable.

### Availability of data and materials

The datasets used and analysed during the current study are available from the corresponding author on reasonable request.

### Clinical trial number

Not applicable.

### Author’s contribution

All authors (A.M.A.A., I.I.Z.I., A.E.H.H., A.I.A.O.) contributed equally to study design, data collection, analysis, manuscript drafting, and critical revisions. All approved the final version.

## Acknowledgement

We are grateful to Samar, Habeeb, Mwada, Badr Aldeen, and Fadwa who assisted in data collection. We appreciate the cooperation and involvement in the study from the orthopaedic patiens, orthopaedic specialiss, and traditional healers.

